# Effect of lifetime exposure to depression on brain structure and function in the UK Biobank

**DOI:** 10.1101/2023.08.31.23294887

**Authors:** Xinyi Wang, Felix Hoffstaedter, Jan Kasper, Simon Eickhoff, Kaustubh R. Patil, Juergen Dukart

## Abstract

**Importance:** Despite several decades of neuroimaging studies reporting brain structural and functional alterations in depression, discrepancies in findings across various studies and limited convergence across several recent meta-analyses have raised questions about the consistency and robustness of the observed brain phenotypes.

**Objective:** To investigate the effects of six different operational criteria of lifetime exposure to depression on functional and structural neuroimaging measures.

**Design, Setting, Participants:** A cross-sectional study analyzed data from the UK biobank in individuals aged 45 to 80 years enrolled from 2014 to 2018. Six operational depression criteria were defined: Help-seeking for depression, Self-reported Depression, Antidepressant usage, Depression defined by Smith, Hospital International Classification of Disease, 10th Edition (ICD-10), and short-form Composite International Diagnostic Interview. Six increasingly conservative groups of lifetime depression were defined based on the six available depression criteria from meeting only one to more restrictive meeting all six criteria. We tested the effect of these definitions on voxel-wise measures of local functional activity, global connectivity, and gray matter volume.

**Main Outcomes and Measures:** Voxel-wise fractional amplitude of low-frequency fluctuations, local connectivity, global connectivity, and gray matter volume.

**Results:** We included 20,484 individuals with lifetime depression (12,645 women [61.73%]; mean [SD] age, 63.92 [7.6] years) and 25,462 healthy individuals (11,384 women [44.7%]; mean [SD] age, 65.05 [7.8] years) from the UK biobank. Across all depression definitions, individuals with lifetime depression displayed regionally consistent decreases in local functional activity in sensorimotor regions but not in global connectivity and gray matter volume. Previous hospital ICD10 diagnosis and antidepressant usage resulted in the most pronounced alterations.

**Conclusions and Relevance:** Lifetime exposure to depression is associated with robust functional changes with more restrictive criteria revealing more pronounced alterations. Different inclusion criteria for depression may strongly contribute to the substantial variation of imaging findings reported in the literature.

## Introduction

Major depressive disorder (MDD) is a common health condition, characterized by low mood, loss of interest or pleasure, feelings of excessive guilt, and thoughts about death or suicide.^1^ The lifetime prevalence of depression is about 11%, with women being more often affected.^2^ Exposure to MDD reduces the quality of life and raises the risk of suicide and self-wounding.^3,4^ Understanding the neurobiological mechanisms underlying MDD is a crucial aspect of developing improved therapeutic options and minimizing negative outcomes.

Numerous smaller functional and structural magnetic resonance imaging (MRI) studies have tested for neurobiological mechanisms of MDD reporting a variety of MDD-related alterations such as hippocampal volume reductions and larger amygdala responses to negative faces.^5–7^ In contrast, neuroimaging meta-analyses and recent large-scale projects revealed less convergent findings with no effects or at most small effect sizes for most of the evaluated neuroimaging modalities.^8–11^ When present, the identified brain alterations did not allow for reliable differentiation between MDD patients and healthy controls with accuracies being only marginally above chance level.^12^ The small effect sizes and lack of differentiation point to currently limited diagnostic value of respective MRI modalities.^12,13^ A confounding factor in this regard which has been largely ignored to date are often-different definitions of depression that have been applied across various cross-sectional and longitudinal studies. Despite this limitation, these studies point to existence of some MDD related brain alterations those being either predisposing or a consequence of MDD diagnosis or treatment. It, however, remains largely unknown if and how far these alterations persist in later life and if prior exposure to depression affects brain function and structure later in life.

Here, we used the UK biobank ^14,15^ (https://www.ukbiobank.ac.uk/) to systematically quantify the magnitude of structural and functional alterations that are associated with lifetime exposure to depression. Making use of the available in-depth phenotyping, we evaluate the effect of different operational criteria of lifetime exposure to depression ranging from self-reports to clinical-defined depression on the magnitude of case-control differences as manifested in brain structure and function.

## Method

### Participants and phenotyping

The present study focused on resting-sate functional fMRI and T1-weight structural images in initial neuroimaging scans of these patients. There were six operational criteria of lifetime exposure to depression which are commonly used in UK biobank depression studies: Help-seeking for depression, Self-reported Depression, Antidepressant usage, Depression defined by Smith,^16^ Hospital International Classification of Disease, 10th Edition (ICD-10), and short-form Composite International Diagnostic Interview (CIDI-SF).^17^ The inclusion criteria of lifetime exposure to depression encompassed meeting one of the following criteria associated with lifetime depression:

1. Help-seeking was responding “Yes” to the following questions: “Have you ever seen a general practitioner for nerves, anxiety, tension, or depression?” [Data-Field: 2090] or “Have you ever seen a psychiatrist for nerves, anxiety, tension, or depression?” [Data-Field: 2100].
2. Self-reported Depression was having experienced depression at present or past [Data-Field ID: 20002] before the neuroimaging scan.
3. The Antidepressant usage was taking antidepressant medication at baseline or follow-up assessment [Data-Field ID: 20003]. The antidepressant codes were listed in Supplementary materials.
4. Depression (Smith) is an approximate measure of lifetime depression by Smith.^16^ Smith et.al defined three types of depression through relevant questions in the mental health questionnaire, including “probable single episode”, “probable mild recurrent”, and “probable severe recurrent”. Participants met one of three lifetime depression were defined as Depression (Smith).
5. Hospital ICD-10 is the hospital recorder for patients’ primary and secondary diagnoses [Data-field ID: 41202 and 41204]. Patients with the diagnosis of the depressive episode (F32-F32.9) or recurrent depressive disorder (F33-F33.8) were included in this criteria.
6. CIDI-SF is derived from the mental health questionnaire.^18^ The CIDI-SF is a brief survey instrument design to identify mental disorders including MDD based on the Diagnostic and Statistical Manual of Mental Disorders (DSM) criteria.^19^

Patients with other lifetime psychiatric criteria were excluded. The detailed exclusion criteria were shown in supplementary methods. To determine whether there are cumulative effects of the above six criteria on brain structural and resting-state functional alterations, we first stratified all patients into six groups according to the number of criteria met. These six graded groups were referred to as “meeting only one criterion” to “meeting all six criteria”. Thereby, “meeting *K* criteria” indicates that participants met exactly *K* criteria rather than at least *K* criteria, in order to keep the six groups without overlapping participants.

Healthy controls (HC) were defined by excluding individuals with indications of psychosis, mental illness, behavior disorder and disease of nervous system. The detailed exclusion criteria of HC were presented in supplementary method. HC groups were defined using two strategies. First, we defined a single HC group which included all individuals who did not meet any of the exclusion criteria (strategy □). Each depression stratum was compared with this HC group after removing unmatched demographics information. Second, we defined six HC groups corresponding to the six depression groups matching them for age, sex and education (strategy II). Strategy I ensures that all patient subgroups are compared to the same, more representative HC group. Strategy II reflects a traditional control group approach to better control for potential demographic confounds. Therefore, we considered the results from the strategy □ (all available healthy individuals) as the primary outcome, while the results from the strategy □ as a control analysis. All participants have provided their informed consent. The UK biobank study was conducted in accordance with the Declaration of Helsinki.

### Imaging data acquisition

MRI data were acquired using a Siemens Skyra 3T scanner (Siemens Healthcare, Erlangen, Germany) using a standard 32-channel head coil, according to a freely available protocol (http://www.fmrib.ox.ac.uk/ukbiobank/protocol/V4_23092014.pdf). As part of the scanning protocol, high-resolution T1-weighted images and resting-state fMRI were obtained. High-resolution T1-weighted images were obtained using an MPRAGE sequence with the following parameters: repetition time (TR) =2000ms, echo time (TE) =2.01ms, 208 slices, flip angle=8°, field of view (FOV) =256mm, matrix=256×256, slice thickness=1.0mm, voxel size=1×1×1mm. The resting-state functional MRI were obtained with following parameters: TR=735ms, TE=39ms, 64 slices, flip angle=52°, FOV=210mm; matrix=88×88; slice thickness=2.4mm, voxel size =2.4×2.4×2.4mm^3^.

### Pre-processing of imaging data

Structural images were preprocessed using SPM12 (Version r7770) (https://www.fil.ion.ucl.ac.uk/spm/software/spm12/) and CAT12 (Version r1720)(https://neuro-jena.github.io/software.html#cat)^20^ with default settings compiled under Matlab 2019b massively parallelized on the JURECA High Performance Computing system (JSC). Raw T1 images were registered to functional images, bias and noise corrected, global intensity normalized, and then segmented into gray matter (GM), white matter, and cerebrospinal fluid. Next, the images were spatially normalized to the standard Montreal Neurological Institute (MNI) templates using Geodesic Shooting.^21^ Relative gray matter volumes (GMV) were computed by dividing the total volume of gray matter by total intracranial volume (TIV). Finally, the GM images were smoothed using an 8-mm full width at half maximum Gaussian kernel. A whole-brain gray matter mask with a probability of gray matter above 0.3 was applied prior to the analyses.

Functional images were preprocessed using SPM12, FSL5.0^22^, and the CONN^23^ toolbox (https://web.conn-toolbox.org/). The functional images were corrected for head motion, normalized for grand-mean intensity. Images were then co-registered to the corresponding high-resolution T1 anatomical images which were transformed into the MNI space. The resulting images were resampled to 3×3×3 mm^3^ voxels, smoothed with a 4-mm-full-width, half-maximum Gaussian kernel. We discarded the first five functional time points to ensure signal equilibrium. Subsequently, temporal band-pass filtering (0.008-0.09 Hz) was performed. Motion parameters (Friston 24 motor parameters)^24^, average white matter and average cerebrospinal fluid signals were regressed out. We then computed commonly applied functional measures using the default settings in the CONN toolbox including the fractional amplitude of low-frequency fluctuations (fALFF), global correlation (GCOR), and local correlation (LCOR). fALFF reflects the amplitude of local low-frequency fluctuations (0.008 Hz to 0.09 Hz) relative to the overall frequency spectrum ^25^. LCOR is computed as the local coherence between a voxel and its neighboring voxels.^26^ GCOR calculates the mean correlation coefficient between BOLD signals of a voxel and all other voxels in the brain.

### Statistical analysis

We performed two-stage analyses to determine brain alterations across increasingly restrictive levels of definitions. Voxel-wise two-sample *t*-tests were conducted in SPM12. First, we compared the voxel-wise fALFF, GCOR, LCOR, and GMV between each stratum of depression and healthy controls (separate for strategy I and strategy □) using *t*-contrasts controlling for age, age squared, sex, and TIV. A whole-brain voxel-wise family-wise error correction at *p*<0.05 was applied for all analyses.

In the second stage, to minimize the impact of different group sizes on the observed differences, we quantified the observed alterations using effect size measures (Cohen’s d). Specifically, we calculated the effect size values between six graded depression and healthy controls based on regions showing significant differences. For this, we employed all available healthy individuals as a single HC group (strategy I). The maps of voxel-wise significant findings from the above *t*-contrasts (separate for increases and decreases) were converted into binary masks to extract the respective effect sizes for all six depression definitions. These masks represent the regions showing significant differences and are further referred to as mask 1 (meeting only one criterion) to mask 6 (meeting all criteria). Theoretically, there were 12 potential masks (6 masks × 2 directional t-contrasts) for each structural or functional measure. Specifically, for each participant we first extracted the individual alterations in respective structural or functional measures in the mask and computed the mean value. We then calculated the effect size between the individuals with different lifetime depression definitions and the single healthy group.

To further explore how the different lifetime depression criteria and their combinations contribute to the observed differences, we computed the effect sizes (Cohen’s d) for group differences for each available combination of depression criteria and each mask. To ensure a more robust estimate of the effect size, we excluded combinations with less than 10 available subjects. To identify which criteria contribute to increases or decreases of the observed effects sizes, we quantified the contribution of each criterion by computing delta effect size (Δeffect size). For this, we categorized all criteria constellations into two groups: one with the specific criterion and the other without the criterion. The Δeffect size was then defined as the mean effect size of combinations with the specific criterion minus the mean effect sizes of combinations without the respective criterion. For each identified mask per modality, this resulted in six Δeffect size values representing contributions of the six criteria.

## Results

### Sample

Of all UK biobank individuals with neuroimaging data in our current application (Application ID: 41655), 20,484 participants met at least one of the criteria of lifetime exposure to depression (12,645 women [61.73%]; mean [SD] age, 63.92 [7.6] years) and 25,462 individuals met the criteria for HC (11,384 women [44.7%]; mean [SD] age, 65.05 [7.8] years). The number of participants admitted in six criteria was: Help-seeking for depression (n=19182), Self-reported Depression (n=4691), Antidepressant Usage (n=4222), Depression (Smith) (n=3166), Hospital ICD-10 (n=1605), CIDI-SF depression (m=3571). For each stratum, the number of participants was: meeting only one criterion (n=10977), meeting two criteria (n=5233), meeting three criteria (n=2574), meeting four criteria (n=1275), meeting five criteria (n=378); meeting all six criteria (n=47). The relevant demographic and clinical information for all groups is provided in Table 1 (all HC, strategy I) and Table S1 (matched HC, strategy □).

**Table 1.** Demographics of the depression group and healthy group (Strategy I) in primary comparison.

|  |  | Depression |  |  | Healthy controls |  |  |
| --- | --- | --- | --- | --- | --- | --- | --- |
| Modality | N | Gender | Age (years) | Education | Gender | Age | Education |
| y | criteria | (female/male |  | (years) | (female/male | (years) | (years) |
|  | met | ) |  |  | ) |  |  |
| Function | 1 | 3936/2572 | 64.01±7.4 | 16.59±3.84 | 6606/7723 | 64.46±7.58 | 16.77±3.7 |
|  | 2 | 1971/1121 | 63.07±7.36 | 16.83±3.7 |  |  |  |
|  | 3 | 1054/530 | 62.74±7.46 | 16.78±3.7 |  |  |  |
|  | 4 | 530/238 | 61.94±7.43 | 16.73±3.8 |  |  |  |
|  | 5 | 174/70 | 61.31±7.76 | 17.44±3.23 |  |  |  |
|  | 6 | 19/10 | 59.82±7.16 | 17.48±2.71 |  |  |  |
| Structure | 1 | 5183/3419 | 63.99±7.38 | 16.58±3.85 | 9263/11070 | 64.48±7.61 | 16.75±3.72 |
|  | 2 | 2628/1482 | 63.09±7.34 | 16.78±3.73 |  |  |  |
|  | 3 | 1377/681 | 62.64±7.38 | 16.82±3.69 |  |  |  |
|  | 4 | 706/335 | 62.03±7.41 | 16.85±3.71 |  |  |  |
|  | 5 | 219/88 | 61.49±7.69 | 17.33±3.32 |  |  |  |
|  | 6 | 27/12 | 59.09±7.06 | 17.77±2.45 |  |  |  |

Group sizes differed slightly for functional and structural analyses due to different drop-outs for quality controls reasons. There were 63 distinct constellations of the six criteria. The number of participants for each combination of the six criteria is shown in Figure 1.

**Figure 1.**
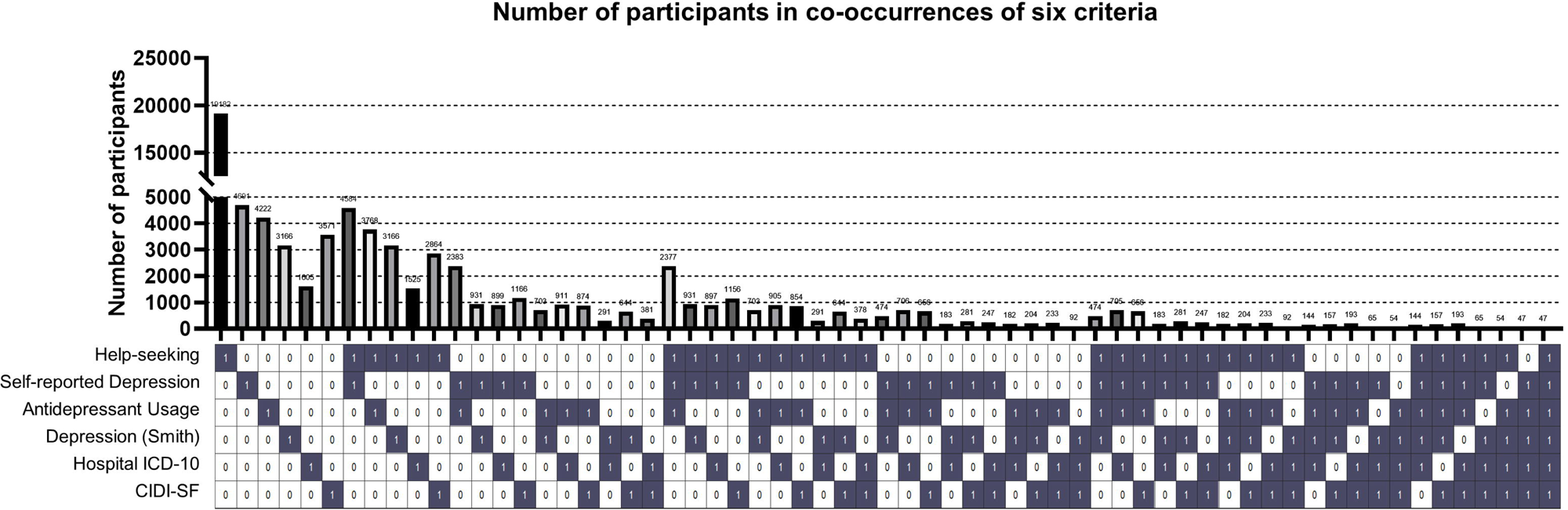
The number of participants in the co-occurrence of six criteria. The first block indicates the number of participants in each constellation. The second block indicates which criteria are involved. The dark element means that the current column contains corresponding criteria.

### Structural and functional alterations

We found significant alterations in all functional and structural measures for most stratums of lifetime depression, except when meeting all six criteria and when meeting only one criterion when using GMV (Figure 2, Figure S1). Group comparisons in functional measures revealed consistently decreased fALFF, GCOR, and LCOR in the lifetime depression groups relative to HC. Clusters of significant functional alterations covered multiple regions encompassing the prefrontal cortex, parietal cortex, middle temporal cortex, fusiform gyrus, occipital cortex, and cerebellum (Table S2-S4). Specifically, fALFF and LCOR alterations displayed similar spatial patterns both showing decreases in the bilateral pre- and postcentral gyrus. Decreased GCOR was primarily observed in the middle inferior temporal cortex, superior temporal cortex, precuneus, insula, and lingual cortex. We found a bidirectional pattern of GMV alterations with increases in the right superior medial frontal cortex and precentral gyrus and decreases in the right hippocampus and superior temporal cortex (Table S5). The outcomes of comparisons of all depression stratums to the respective matched HC groups were largely consistent with the primary analysis (Table S6-S9).

**Figure 2.**
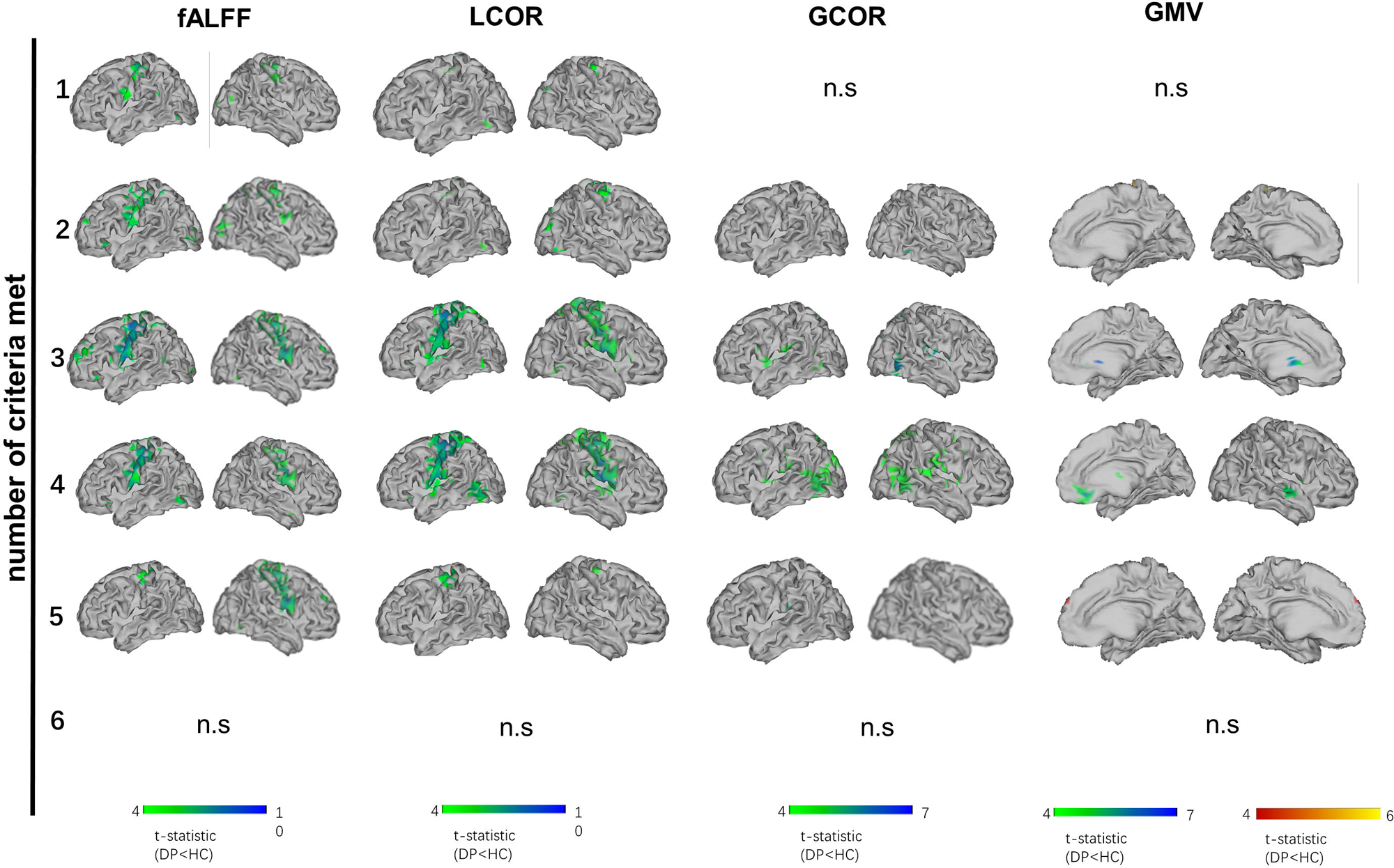
Clusters of significant functional or structural differences between different depression stratums and healthy controls. The n.s indicates no significant clusters. Abbreviations: fALFF: fractional amplitude of low-frequency fluctuations. GCOR: global correlation, LCOR: local correlation, GMV: gray matter volumes.

### Group differences between lifetime depression and HC across increasingly conservative depression

As some of the observed imaging differences across the increasingly conservative depression definitions may simply reflect differences in sample size, we next computed effect sizes for each constellation of depression and each significance mask derived from the above group-comparisons (Figure 3). Sixty-three different constellations of depression criteria were available in the UK biobank (Figure 3B). Effect sizes increased with an increased number of criteria met (Figure 3A). Effect sizes were negative for all functional measures. The “meeting all 6 criteria” definition consistently showed largest effect sizes in fALFF, LCOR and GCOR. Effect sizes were generally lower for GMV and only masks 3 and masks 4 from the decreased GMV displayed consistently negative but overall weak effect sizes. Results for masks 2 and 4 that were derived from the increased GMV clusters were not consistent.

**Figure 3.**
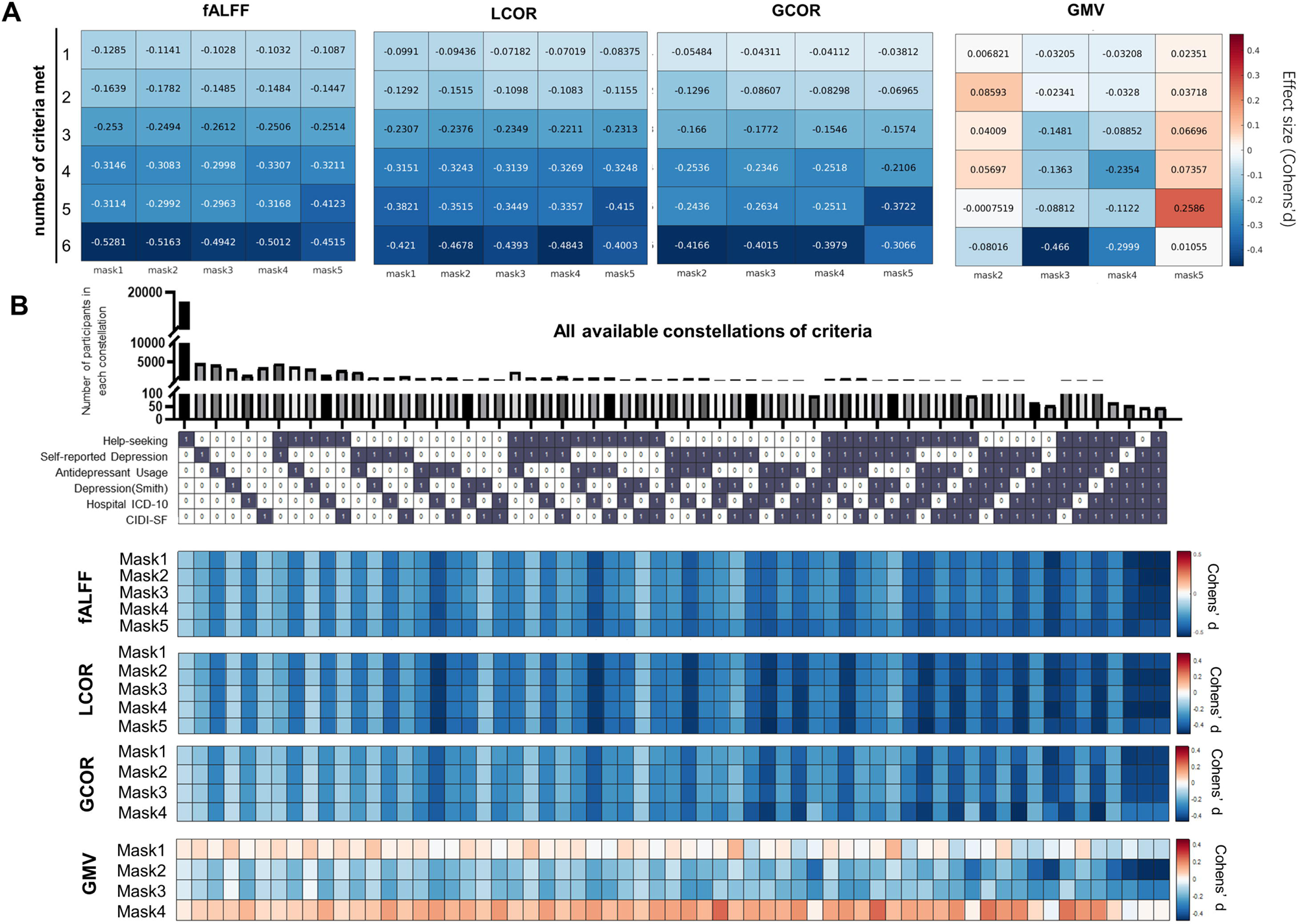
The impact of different lifetime depression on the observed imaging alterations. (A) The impact of each stratum of lifetime depression across all masks. The rows indicate six lifetime depression stratums and columns indicate masks. The masks show significant differences between each depression group and healthy controls (mask 1 – meeting one criterion to mask 5 – meeting five criteria). The effect sizes are calculated in all depression stratum–identified mask pairs. (B) The impact of all available constellations of lifetime depression across all masks. The rows indicate observed masks and columns indicate all available constellations/combinations of six criteria. Red indicates a positive effect size value (increased imaging measures in depression), while blue indicates a negative effect size value (decreased imaging measures in healthy controls). Abbreviations: fALFF: fractional amplitude of low-frequency fluctuations. GCOR: global correlation, LCOR: local correlation, GMV: gray matter volumes.

### Contribution of six criteria to the observed alterations

Next, we aimed to quantify how the different criteria contribute to the observed group differences. Higher (negative and positive) Δeffect size values indicate stronger contribution of the respective criterion. Constellations involving Hospital ICD-10 and antidepressants antidepressant displayed consistently strongest contributions to the observed effect sizes for all functional measures and for the GMV decrease mask (Figure 4A, B). None of the criteria displayed a consistent contribution for the GMV increase mask.

**Figure 4.**
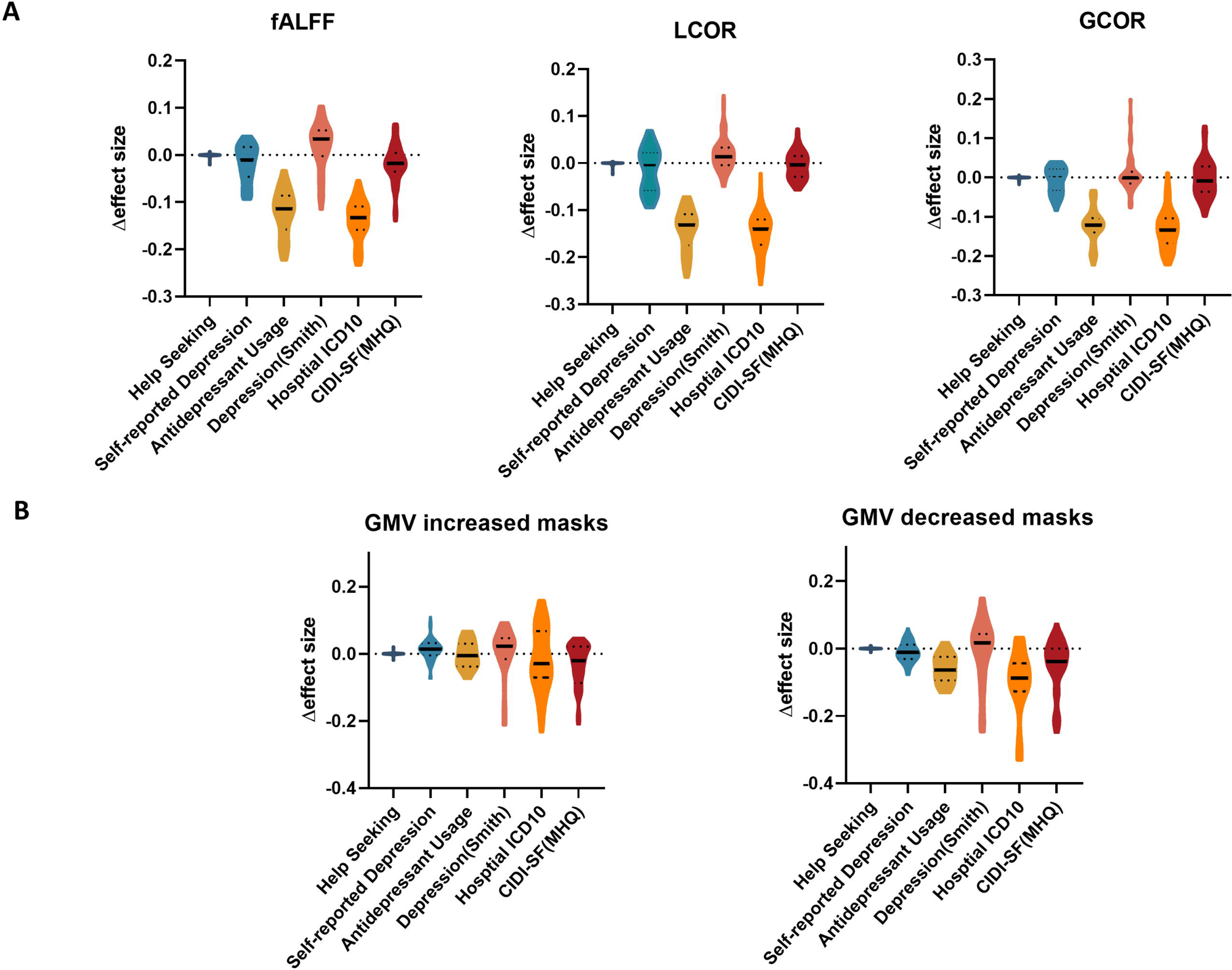
Violin plots to show contribution of six criteria to the observed alterations. The distribution of each criterion in Δeffect size was presented in each violin. The extent of the violin plot is the range of Δeffect size. The inside solid line indicates the median of the Δeffect size, while dot lines indicate the quartiles. Each side of violin plot is the kernel density estimation to show the distribution shape of the Δeffect size. (A) The contributions (Δeffect size) of each criterion in fALFF, LCOR, GCOR. (C) The contributions (Δeffect size) of each criterion in increased (mask1, mask4) and decreased masks (mask2, mask3) for GMV.

## Discussion

Here, we systematically explored the effects of lifetime exposure to depression on established brain structural and functional measures. We find both functional and structural alterations to strongly vary with different definitions of depression, as available in the UK biobank, with stricter definitions resulting in stronger functional but not necessarily structural alterations. In particular, decreases in local functional activity were consistent across all definitions with Hospital ICD-10 diagnosis and antidepressant usage contributing strongest to the observed alterations.

Increases as well as decreases in brain functional measures have been associated with depression in previous studies.^6,27–29^ Contrasting some of these findings, we find only decreases of local activity and local and global connectivity measures across all constellations of lifetime depression. These findings are in line with recent larger studies and meta-analyses reporting reduced functional connectivity in the default mode networks and convergent regions only for decreased functional findings in MDD.^9,30^ Importantly, despite the matching direction of change we observe only limited convergence in sensorimotor regions with these studies in terms of location of the observed alteration patterns. Whilst differences in the samples and applied methodology likely account for some of the discrepancies, a main difference is also in the characterization of the lifetime depression criterion in our study as opposed to the more acute effects of depression reported in the literature. Our findings suggest the existence of a persistent depression-related brain functional phenotype with small to moderate effect sizes.

We find antidepressant medication and the ICD-10 depression diagnosis to be the strongest contributing criteria to the observed functional alterations. Only limited contribution was observed for all other applied criteria. As the anti-depressant medication status is typically a direct consequence of a depression diagnosis only limited conclusions can be made with respect to any causal interpretation of the observed alterations being the cause or consequence of requiring medication. Despite this limitation, as there are numerous treatment interventions for depression that are not medication-based, i.e. psychological (e.g., cognitive behavioral therapy and stimulation therapies) and as patients often refuse antidepressant treatment, these two criteria are not necessarily identical. The separate contributions of ICD-10 diagnosis and antidepressant usage observed in our study may point to such a differential contribution of both variables to the observed decreases. A causal interpretation is limited also in this case as the observed effects still be attributed to either the necessity of prescribing anti-depressants or the consequence of being exposed to such.

We find decreased local functional activity and synchronicity in pre- and postcentral gyrus and decreased global functional connectivity in parts of the limbic system to be consistently associated with lifetime depression. The decreased local activity in sensorimotor regions may be attributed to consequence of being exposure to treatments and depression-associated vulnerability. For treatment effects, a meta-analysis found that stimulation therapy for depression altered the activity in the right precentral gyrus, right posterior cingulate, left inferior frontal gyrus, and left middle frontal gyrus.^31^ After electroconvulsive therapy, fALFF was reported to decrease in the right precentral gyrus. ^32^ Additionally, antidepressant resulted in decrease in hyperconnectivity within the limbic system.^33^ Contrary to the functional measures, the findings for GMV are only partially consistent to previous studies. The observed reduced hippocampal volume is commonly reported in meta-analyses and case-control studies.^34^ The effects size of GVM alternations was substantially smaller as compared to functional measures.

Overall, our findings provide clear evidence for the impact of different depression definitions on the observed imaging outcomes. As many other cohorts, the UK biobank contains extensive data items associated with depression. However, the presence of multiple sources of information also presents challenges in achieving consistent definitions of depression across different studies. As stricter definitions often conflict with the available sample size, researchers might be tempted to apply fewer restrictions to putatively increase statistical power. Here we show that such an approach may become futile in case of neuroimaging as less restrictive definitions may lead to dilution of potential imaging alterations. Recent studies have explored the effects of different depression phenotypes in the UK Biobank on genetic and cortical thickness measures.^35^ Whilst one of these studies suggested that a broader definition of depression may provide more tractable phenotypes,^36^ others recommended more restrictive definitions suggesting that minimal phenotyping may bias potential findings by introducing conceptual differences in the selected cohorts.^37,38^ Extending on these findings, we show that while restrictiveness is generally rather beneficial, the specific criteria should be carefully weighted and evaluated. Only two of the six applied depression definition criteria consistently contributed to the magnitude of the functional imaging alterations observed in our study. These findings suggest that a mere restrictiveness may become counter-productive and that the impact of each criterion needs to be more carefully evaluated in future research.

In conclusion, our study demonstrates the significant influence of different definitions of lifetime depression on functional and structural imaging measures. We evaluated six depressive criteria, ranging from minimal to more restrictive clinically-defined criteria. We identified specific criteria associated with stronger differences in imaging phenotypes compared with control groups and provide recommendations for selecting appropriate criteria in future research endeavors.

### Data Sharing Statement

The images and phenotypic data that support the current findings are available from UK Biobank. Data would be available from the authors upon reasonable application and with permission of UK Biobank (http://www.ukbiobank.ac.uk/).

## Acknowledge

The authors gratefully acknowledge the computing time granted by the JARA Vergabegremium and provided on the JARA Partition part of the supercomputer JURECA at Research Centre Jülich.

