## Supplementary Materials for "Effect of lifetime exposure to depression on brain structure and function in the UK Biobank"

**The exclusion criteria for lifetime depression**

The exclusion criteria of individuals in the lifetime depression group were:

(1) Having self-reported psychosis. The self-reported psychosis was defined as having experienced schizophrenia or mania/bipolar disorder/manic depression at present or past [Data-Field ID: 20002].
(2) Having taken antipsychotic medications. The antipsychotic codes were listed in supplementary materials.
(3) The hospital records [Data-Field ID: 41202 or 41204] included Schizophrenia, schizotypal and delusional disorders (F20-F29), and other mood disorders (F30-F39, excluding depression code: F32-F33).
(4) Having been defined as psychosis in mental health questionnaire (MHQ) screen. The answer to the question “Have you been diagnosed with one or more of the following mental health problems by a professional, even if you don’t have it currently” [Data-field ID: 20544] is “Schizophrenia” or “Any other type of psychosis or psychotic illness”, or “Mania, hypomania, bipolar or manic-depression”.

**The exclusion criterion for healthy controls**

The healthy individuals were enrolled with the following exclusive criterion:

(1) Did not meet the criteria for any indications of depression as described in the above depression phenotypes.
(2) Did not meet the criteria for any indications of psychosis which was described in the depression exclusive criterion, including self-reported psychosis, antipsychotic medications usage, MHQ psychosis.
(3) Did not endorse disorder in any mental illness and behavior disorder (Hospital ICD10 Chapter V, F00-F99) and diseases of the nervous system (Hospital ICD10 Chapter VI G00-G99）.

**Medications codes**

The antidepressants are coded as follows [UKB Data-Coding 4]:

1140879616, 1140921600, 1140879540, 1140867878, 1140916282, 1140909806, 1140867888, 1141152732, 1141180212, 1140879634, 1140867876, 1140882236, 1141190158, 1141200564, 1140867726, 1140879620, 1140867818, 1140879630, 1140879628, 1141151946, 1140867948, 1140867624, 1140867756, 1140867884, 1141151978, 1141152736, 1141201834, 1140867690, 1140867640, 1140867920, 1140867850, 1140879544, 1141200570, 1140867934, 1140867758, 1140867914, 1140867820, 1141151982, 1140882244, 1140879556, 1140867852, 1140867860, 1140917460, 1140867938, 1140867856, 1140867922, 1140910820, 1140882312, 1140867944, 1140867784, 1140867812, 1140867668.

The antipsychotics in UK biobank are coded as follows:

1140868170 1140928916 1141152848 1140867444 1140879658 1140868120 1141153490 1140867304 1141152860 1140867168 1141195974 1140867244 1140867152 1140909800 1140867420 1140879746 1141177762 1140867456 1140867952 1140867150 1141167976 1140882100 1140867342 1140863416 1141202024 1140882098 1140867184 1140867092 1140882320 1140910358 1140867208 1140909802 1140867134 1140867306 1140867210 1140867398 1140867078 1140867218 1141201792 1141200458 1140867136 1140879750 1140867180 1140867546 1140928260 1140927956.

**Supplementary Figures**

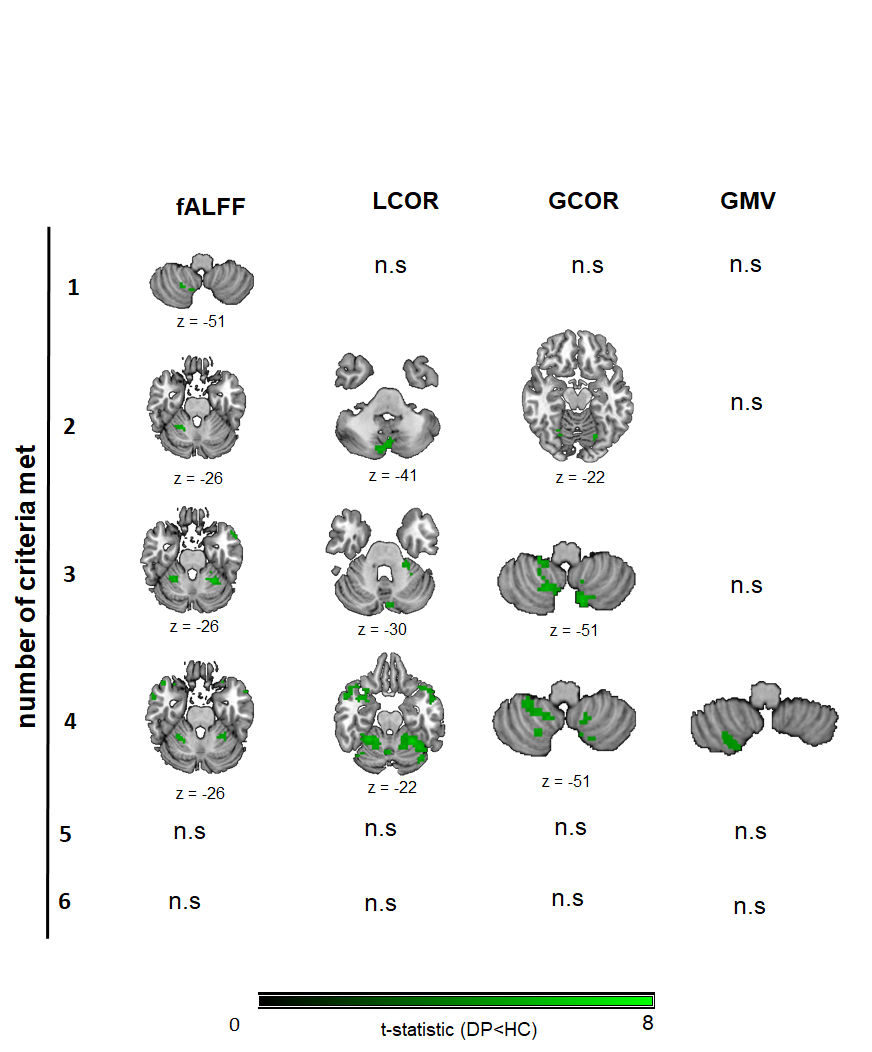

Figure S1. The cerebellum parts of significant clusters. The n.s indicates the no significant difference in the cerebellum. Abbreviations: fALFF: fractional amplitude of low-frequency fluctuations. GCOR: global correlation, LCOR: local correlation, GMV: gray matter volumes.

**Supplementary Tables**

Table S1 Demographic of six graded depression involving analysis

| Modality | N criteria met | Gender (female/male) | Age (years) | Education (years) |
| --- | --- | --- | --- | --- |
| Function | 1 | 3936/2572 | 64.01±7.4 | 16.59±3.84 |
|  | 2 | 1971/1121 | 63.07±7.36 | 16.83±3.7 |
|  | 3 | 1054/530 | 62.74±7.46 | 16.78±3.7 |
|  | 4 | 530/238 | 61.94±7.43 | 16.73±3.8 |
|  | 5 | 174/70 | 61.31±7.76 | 17.44±3.23 |
|  | 6 | 19/10 | 59.82±7.16 | 17.48±2.71 |
| Structure | 1 | 5183/3419 | 63.99±7.38 | 16.58±3.85 |
|  | 2 | 2628/1482 | 63.09±7.34 | 16.78±3.73 |
|  | 3 | 1377/681 | 62.64±7.38 | 16.82±3.69 |
|  | 4 | 706/335 | 62.03±7.41 | 16.85±3.71 |
|  | 5 | 219/88 | 61.49±7.69 | 17.33±3.32 |
|  | 6 | 27/12 | 59.09±7.06 | 17.77±2.45 |

Table S1 Demographic of the single healthy groups (Strategy Ⅱ)

|  | Gender (female/male) | Age (years) | Education (years) |
| --- | --- | --- | --- |
| Function | 6606/7723 | 64.46±7.58 | 16.77±3.7 |
| Structure | 9263/11070 | 64.48±7.61 | 16.75±3.72 |

Table S1 Demographic of six matched healthy groups (Strategy Ⅱ)

| Modalities |  | Healthy controls | | |
| --- | --- | --- | --- | --- |
|  | N criteria met | Gender (female/male) | Age (mean±std) | Years of Education (mean±std) |
| Function | 1 | 3945/2573 | 63.76±7.39 | 16.8±3.69 |
|  | 2 | 1976/1120 | 63.05±7.31 | 16.87±3.68 |
|  | 3 | 1053/534 | 62.66±7.44 | 16.86±3.61 |
|  | 4 | 529/236 | 61.93±7.44 | 16.98±3.56 |
|  | 5 | 172/71 | 61.39±7.4 | 17.59±2.97 |
|  | 6 | 19/10 | 59.85±7.2 | 17.52±2.43 |
| Structure | 1 | 5184/3418 | 63.89±7.41 | 16.68±3.79 |
|  | 2 | 2630/1480 | 63.13±7.32 | 16.91±3.63 |
|  | 3 | 1375/683 | 62.63±7.36 | 16.9±3.59 |
|  | 4 | 707/334 | 62.06±7.38 | 16.9±3.64 |
|  | 5 | 220/87 | 61.6±7.62 | 17.36±3.23 |
|  | 6 | 28/11 | 59.49±6.81 | 17.9±2.19 |

Table S2. Regions showing significant differences in fALFF between individuals with lifetime depression and the single HC group (Strategy I)

| N criteria me | Contrast | Cluster size | Cluster p-value | Peak T statistic | Peak effect size | MNI coordinates | Anatomical region |
| --- | --- | --- | --- | --- | --- | --- | --- |
| 1 | DP<HC | 43 | p<0.001 | 6.440 | -0.109 | [-51;27;-12] | left posterior orbital gyrus |
|  |  | 34 | p<0.001 | 6.407 | -0.097 | [-21;-84;45] | left superior temporal cortex |
|  |  | 31 | p<0.001 | 6.373 | -0.097 | [-63;-51;24] | left supraMarginal gyrus and middle temproal cortex |
|  |  | 49 | p<0.001 | 6.142 | -0.103 | [24;-48;-27] | right Lobule IV, V, VI of cerebellar hemisphere |
| 2 | DP<HC | 55 | p<0.001 | 6.208 | -0.134 | [48;-69;-12] | right inferior temporal and occipital cortex |
|  |  | 156 | p<0.001 | 6.181 | -0.139 | [-33;48;33] | left middle and superior frontal cortex |
|  |  | 49 | p<0.001 | 6.027 | -0.133 | [21;-51;-27] | righ Lobule IV, V, VI of cerebellar hemisphere |
|  |  | 69 | p<0.001 | 5.969 | -0.150 | [-21;-57;-48] | left Lobule VIIB, VIII of cerebellar hemisphere |
| 3 | DP<HC | 5545 | p<0.001 | 9.552 | -0.250 | [-42;-33;54] | bilateral medial and lateral pre- and postercentral gyrus |
|  |  | 271 | p<0.001 | 7.546 | -0.220 | [15;-60;-51] | bilateral loublue VIII and IX of cerebellar |
|  |  | 712 | p<0.001 | 7.494 | -0.220 | [-36;48;30] | left superiror medial frontal cortex |
|  |  | 318 | p<0.001 | 6.671 | -0.205 | [27;42;27] | right superior and medial frontal cortex |
|  |  | 156 | p<0.001 | 6.580 | -0.222 | [36;30;-12] | right inferior frontal gyrus |
|  |  | 63 | p<0.001 | 6.463 | -0.198 | [-21;-39;-24] | Left Lobule IV, V,VI of cerebellar hemisphere |
|  |  | 161 | p<0.001 | 6.362 | -0.167 | [-18;-102;15] | left middle and superiror occipital cortex |
|  |  | 90 | p<0.001 | 6.303 | -0.191 | [21;-51;-24] | right Lobule IV, V,VI of cerebellar hemisphere |
|  |  | 56 | p<0.001 | 6.168 | -0.171 | [48;-66;-12] | right inferior temporal gyrus |
|  |  | 92 | p<0.001 | 6.135 | -0.191 | [-57;-63;12] | left middle temporal gyrus |
|  |  | 47 | p<0.001 | 6.069 | -0.174 | [3;54;0] | right superior and medial frontal cortex |
|  |  | 29 | p<0.001 | 5.817 | -0.149 | [21;-96;24] | right occipital cortex |
|  |  | 25 | p<0.001 | 5.615 | -0.162 | [-21;-66;57] | left superior parietal cortex |
| 4 | DP<HC | 4012 | p<0.001 | 9.061 | -0.317 | [-3;-9;51] | bilateral pre- and postcentral gyrus |
|  |  | 79 | p<0.001 | 7.045 | -0.238 | [-12;-99;24] | left superior and middle occipital cortex |
|  |  | 198 | p<0.001 | 6.419 | -0.279 | [-45;-66;-9] | left middle and inferior occipital cortex |
|  |  | 74 | p<0.001 | 6.411 | -0.265 | [24;-51;-21] | right Lobule IV, V,VI of cerebellar hemisphere |
|  |  | 60 | p<0.001 | 6.357 | -0.239 | [-9;66;24] | left superior frontal cortex |
|  |  | 57 | p<0.001 | 6.331 | -0.276 | [-6;-75;-45] | left Lobule VII, VIII of cerebellar hemisphere |
|  |  | 30 | p<0.001 | 6.327 | -0.215 | [12;-99;21] | right superior occipital cortex |
|  |  | 55 | p<0.001 | 6.216 | -0.237 | [39;57;15] | right superior and middle frontal cortex |
|  |  | 48 | p<0.001 | 6.162 | -0.285 | [-24;0;-12] | left amygdala and putamen and putamen |
|  |  | 36 | p<0.001 | 6.058 | -0.250 | [12;-69;-48] | right Lobule VIII of cerebellar hemisphere |
|  |  | 33 | p<0.001 | 6.006 | -0.265 | [24;3;-12] | right amygdala |
|  |  | 44 | p<0.001 | 5.821 | -0.271 | [36;21;-33] | right superior and middle temporal cortex |
|  |  | 38 | p<0.001 | 5.738 | -0.245 | [-18;-54;-18] | left Lobule IV, V of cerebellar hemisphere |
|  |  | 27 | p<0.001 | 5.568 | -0.240 | [-48;21;-15] | left posterior orbital gyrus and superior tempral pole cortex |
|  |  | 39 | p<0.001 | 5.511 | -0.226 | [45;-69;-9] | right fusiform |
|  |  | 28 | p<0.001 | 5.465 | -0.243 | [-57;9;-24] | left middle temporal cortex |
| 5 | DP<HC | 312 | p<0.001 | 6.700 | -0.410 | [-42;-36;63] | left pre-and postcentral gyrus |

Table S3 Regions showing significant differences in LCOR between individuals with lifetime depression and the single HC group (Strategy I)

| N criteria met | Contrast | Cluster size | Cluster p-value | Peak T statistic | Peak effect size | MNI coordinates | Anatomical region |
| --- | --- | --- | --- | --- | --- | --- | --- |
| 1 | DP<HC | 230 | p<0.001 | 6.686 | -0.088 | [0;-18;72] | left paracentral loubel and support motor area |
|  |  | 299 | p<0.001 | 6.329 | -0.086 | [39;-18;66] | right pre- and postcentral gyrus |
|  |  | 258 | p<0.001 | 6.216 | -0.087 | [-48;-27;60] | left pre-and postcentral gyrus |
|  |  | 34 | p<0.001 | 6.032 | -0.084 | [-9;-102;18] | left superior occipital cortex |
|  |  | 112 | p<0.001 | 5.487 | -0.085 | [36;-84;27] | right middle occipital cortex |
|  |  | 41 | p<0.001 | 5.281 | -0.083 | [-51;-72;-9] | left inferior temporal cortex |
| 2 | DP<HC | 1155 | p<0.001 | 6.990 | -0.129 | [30;-15;63] | right pre- and postcentral gyrus |
|  |  | 82 | p<0.001 | 6.232 | -0.130 | [3;-75;-42] | left Crus II, Lobule VIII of cerebellar hemisphere |
|  |  | 87 | p<0.001 | 6.036 | -0.124 | [48;-66;-15] | right inferior occipital and temporal cortex |
|  |  | 122 | p<0.001 | 5.996 | -0.131 | [12;-57;-51] | right Lobule VIII, Ⅸ, Ⅹ of cerebellar hemisphere |
|  |  | 356 | p<0.001 | 5.991 | -0.123 | [-33;-21;63] | left pre-and postcentral gyrus |
|  |  | 154 | p<0.001 | 5.830 | -0.121 | [6;-96;18] | left middle and superior occipital cortex |
|  |  | 175 | p<0.001 | 5.826 | -0.110 | [30;-87;33] | right superior and middle occipital cortex |
|  |  | 86 | p<0.001 | 5.629 | -0.119 | [-54;-75;3] | left inferior occipital and temporal cortex |
|  |  | 27 | p<0.001 | 5.605 | -0.110 | [-48;33;30] | left triangular part of inferior frontal cortex |
|  |  | 30 | p<0.001 | 5.309 | -0.102 | [57;-12;36] | right pre-and postcentral gyrus |
| 3 | DP<HC | 6979 | p<0.001 | 9.066 | -0.228 | [-45;-33;63] | bilateral pre- and postcentral gyrus |
|  |  | 594 | p<0.001 | 6.922 | -0.203 | [3;-72;-42] | bilateral Lobule VIII and IX of cerebellar heimsphere |
|  |  | 124 | p<0.001 | 6.619 | -0.174 | [-21;-48;-15] | left fusiform gyrus |
|  |  | 105 | p<0.001 | 5.984 | -0.159 | [48;-63;-12] | right inferior temporal and occipital cortex |
|  |  | 60 | p<0.001 | 5.507 | -0.155 | [63;-57;9] | right middle temporal cortex |
|  |  | 105 | p<0.001 | 5.479 | -0.160 | [-33;30;0] | left triangular part of inferior frontal gyrus |
|  |  | 87 | p<0.001 | 5.463 | -0.156 | [-48;-66;-6] | left inferior temporal and occipital cortex |
|  |  | 41 | p<0.001 | 5.230 | -0.144 | [-54;-60;12] | left middle temporal gyrus |
|  |  | 52 | p<0.001 | 5.163 | -0.161 | [15;-36;-3] | right fusiform gyrus and lingual cortex |
| 4 | DP<HC | 7633 | p<0.001 | 9.593 | -0.313 | [-48;-27;60] | bilateral pre-and postcentral gyrus |
|  |  | 1212 | p<0.001 | 7.797 | -0.300 | [-48;-69;-6] | left middle and inferior temproal cortex and bilateral fusiform gyrus |
|  |  | 423 | p<0.001 | 6.627 | -0.264 | [-3;-75;-42] | bilateral Lobule VIIB, and VIII of cerebellar hemisphere |
|  |  | 37 | p<0.001 | 5.891 | -0.217 | [-42;33;39] | left middle frontal cortex |
|  |  | 61 | p<0.001 | 5.707 | -0.205 | [-18;-99;21] | left superior occipital cortex |
|  |  | 29 | p<0.001 | 5.656 | -0.216 | [-24;0;-9] | left putamen and amygdala |
|  |  | 36 | p<0.001 | 5.618 | -0.211 | [42;27;36] | right middle frontal cortex |
|  |  | 38 | p<0.001 | 5.591 | -0.205 | [-6;39;24] | left superior medial frontal cortex and anterior cingulate cortex |
|  |  | 47 | p<0.001 | 5.444 | -0.208 | [39;-69;-48] | right Crus II, Lobule VIIB, and VIII of cerebellar hemisphere |
| 5 | DP<HC | 464 | p<0.001 | 6.892 | -0.394 | [-45;-27;63] | left pre-and postcentral gyrus |
|  |  | 48 | p<0.001 | 6.043 | -0.392 | [-48;-21;21] | left ralandic operculum and supramarginal cortex |
|  |  | 193 | p<0.001 | 5.770 | -0.384 | [-3;-18;51] | bilateral support motor area |
|  |  | 46 | p<0.001 | 5.691 | -0.360 | [-63;-6;12] | left postcentral gyrus and superior temporal cortex |
|  |  | 255 | p<0.001 | 5.664 | -0.361 | [39;-30;48] | right pre- and postcentral gyrus |
|  |  | 106 | p<0.001 | 5.655 | -0.363 | [-12;-33;78] | bilateral paracentral lobule; bilateral pre- and postcentral cortex |
|  |  | 59 | p<0.001 | 5.357 | -0.361 | [66;-12;27] | Right postcentral gyrus |

Table S4 Regions showing significant differences in GCOR between individuals with lifetime depression and the single HC group (Strategy I)

| N criteria met | Contrast | Cluster size | Cluster p-value | Peak T statistic | Peak effect size | MNI coordinates | Anatomical region |
| --- | --- | --- | --- | --- | --- | --- | --- |
| 2 | DP<HC | 28 | p<0.001 | 6.051 | -0.113 | [-54;-75;3] | left middle and inferior temporal cortex |
|  |  | 41 | p<0.001 | 5.850 | -0.114 | [57;-63;-3] | right middle and inferior temporal cortex |
|  |  | 33 | p<0.001 | 5.771 | -0.111 | [18;-54;-9] | right lingual and fusiform gyrus |
|  |  | 26 | p<0.001 | 5.457 | -0.109 | [-39;21;30] | left inferior frontal cortex |
| 3 | DP<HC | 826 | p<0.001 | 7.483 | -0.170 | [-39;0;12] | left insula and superior temporal cortex |
|  |  | 2852 | p<0.001 | 7.071 | -0.175 | [-21;-45;-15] | left middle cingulate cortex, precuneus and lingual cortex |
|  |  | 669 | p<0.001 | 6.245 | -0.162 | [39;-3;15] | right insular and rolandic operculum |
|  |  | 192 | p<0.001 | 6.225 | -0.182 | [9;-69;-45] | bilateral Lobule VIII of cerebellar hemisphere |
|  |  | 43 | p<0.001 | 5.675 | -0.150 | [48;3;39] | right precentral cortex |
|  |  | 90 | p<0.001 | 5.408 | -0.149 | [-60;-18;42] | left inferior parietal superior cortex |
|  |  | 36 | p<0.001 | 5.283 | -0.160 | [39;6;-15] | right superior temporal cortex |
|  |  | 31 | p<0.001 | 5.225 | -0.140 | [-48;3;39] | left precentral cortex |
| 4 | DP<HC | 7471 | p<0.001 | 7.684 | -0.250 | [9;-60;69] | bilateral precuneus, lingual cortex, fusiform, middle and superior temporal cortex |
|  |  | 148 | p<0.001 | 6.912 | -0.231 | [-36;3;33] | left precentral cortex |
|  |  | 191 | p<0.001 | 6.772 | -0.249 | [15;-66;-45] | bilateral Lobule VIII and left Lobule VIIB of cerebellar hemisphere |
|  |  | 32 | p<0.001 | 6.419 | -0.236 | [-24;0;-12] | left amygdala and putamen |
|  |  | 52 | p<0.001 | 6.238 | -0.241 | [27;-42;-48] | right Lobule VIII of cerebellar hemisphere |
|  |  | 165 | p<0.001 | 5.824 | -0.224 | [39;9;33] | right precentral cortex and inferior frontal cortex |
|  |  | 28 | p<0.001 | 5.355 | -0.199 | [-33;-42;42] | left inferior parietal cortex |
|  |  | 25 | p<0.001 | 5.121 | -0.198 | [-36;-3;54] | left precentral cortex |
| 5 | DP<HC | 59 | p<0.001 | 5.825 | -0.361 | [-48;-33;24] | left supramarginal gyrus and superior temproal cortex |

Table S5 Regions showing significant differences in GMV between individuals with lifetime depression and the single HC group (Strategy I)

| N criteria met | | Contrast | Cluster size | Cluster p-value | Peak T statistic | Peak effect size | MNI coordinates | Anatomical region |
| --- | --- | --- | --- | --- | --- | --- | --- | --- |
|  | 2 | DP>HC | 56 | p<0.001 | 5.286 | 0.086 | [12;-30;75] | right pre- and post-central gyrus |
|  | 3 | DP<HC | 396 | p<0.001 | 6.855 | -0.142 | [3;4.5;3] | bilateral Olfactory cortex |
|  |  | DP<HC | 60 | p<0.001 | 5.038 | -0.110 | [18;-13.5;-10.5] | right hippocampus |
|  | 4 | DP<HC | 1608 | p<0.001 | 6.285 | -0.183 | [0;33;-15] | bilateral medial frontal cortex |
|  |  |  | 218 | p<0.001 | 6.058 | -0.185 | [28.5;30;36] | right superior and medial frontal cortex |
|  |  |  | 1051 | p<0.001 | 5.761 | -0.179 | [58.5;-10.5;-9] | right superior and middle temporal cortex |
|  |  |  | 434 | p<0.001 | 5.568 | -0.182 | [-3;-16.5;49.5] | bilateral middle cingulate cortex |
|  |  |  | 147 | p<0.001 | 5.317 | -0.162 | [18;-16.5;-12] | right hippocampus |
|  |  |  | 70 | p<0.001 | 5.192 | -0.160 | [21;-45;-4.] | right lingual cortex and parahippocampus |
|  |  |  | 161 | p<0.001 | 5.129 | -0.162 | [-49.5;3;-3] | left superior temporal cortex |
|  |  |  | 391 | p<0.001 | 5.074 | -0.176 | [-49.5;3;-28.5] | left inferior and middle temporal cortex |
|  |  |  | 208 | p<0.001 | 5.036 | -0.157 | [0;-9;9] | bilateral media dorsal medial magnocellular |
|  |  |  | 113 | p<0.001 | 4.981 | -0.163 | [-45;-28.5;48] | left postcentral gyrus |
|  |  |  | 171 | p<0.001 | 4.921 | -0.163 | [1.5;37.5;25.5] | left anterior cingulate cortex |
|  |  |  | 84 | p<0.001 | 4.920 | -0.158 | [-58.5;-33;16.5] | left superior temporal |
|  |  |  | 112 | p<0.001 | 4.885 | -0.160 | [52.5; 3;0] | right insula and righ rolandic operculum |
|  |  |  | 215 | p<0.001 | 4.870 | -0.153 | [25.5;-63;-58.5] | right Lobule VIII of cerebellar hemisphere |
|  |  |  | 177 | p<0.001 | 4.856 | -0.157 | [43.5;-15;9] | right insula |
|  |  |  | 51 | p<0.001 | 4.714 | -0.150 | [30;12;-25.5] | right temporal pole: superior temporal gyrus |
|  | 5 | DP>HC | 62 | p<0.001 | 4.670 | 0.259 | [12;64.5;33] | right superior medial frontal cortex |

Table S6. Regions showing significant differences in fALFF between individuals with lifetime depression and matched HC group (Strategy Ⅱ)

| N criteria met | Contrast | Cluster size | Cluster p-value | Peak T statistic | MNI coordinates | Anatomical region |
| --- | --- | --- | --- | --- | --- | --- |
| 2 | DP<HC | 537 | p<0.001 | 6.61 | [27;-78;-6] | right Fusiform gyrus, lingual gyrus, occipital pole |
|  |  | 196 | p<0.001 | 5.91 | [-39;-81;-9] | left middle occipital cortex |
|  |  | 75 | p<0.001 | 5.72 | [-6;-21;66] | left thalamus |
|  |  | 46 | p<0.001 | 5.65 | [-30;-9;60] | left precentral gyrus |
| 3 | DP<HC | 2759 | p<0.001 | 8.05 | [0;-30;63] | bilateral medial and lateral primary sensorimotor regions |
|  |  | 90 | p<0.001 | 6.46 | [18;-60;-51] | right Lobule VIII of cerebellar hemisphere |
|  |  | 121 | p<0.001 | 6.17 | [-39;-24;21] | left parietal operculum cortex |
|  |  | 70 | p<0.001 | 6.04 | [0;60;27] | superior medial frontal |
|  |  | 115 | p<0.001 | 5.96 | [-27;60;21] | left superior frontal cortex |
|  |  | 32 | p<0.001 | 5.93 | [-30;-51;-48] | left Lobule VIII of cerebellar hemisphere |
|  |  | 28 | p<0.001 | 5.79 | [0;54;-9] | left medial frontal cortex |
|  |  | 34 | p<0.001 | 5.72 | [-9;-72;-48] | left Lobule VIII of cerebellar hemisphere |
|  |  | 30 | p<0.001 | 5.57 | [42;-69;-12] | right lateral occipital cortex |
|  |  | 30 | p<0.001 | 5.55 | [-27;-87; 0] | left middle occipital cortex |
| 4 | DP<HC | 137 | p<0.001 | 7.63 | [0;-9;51] | bilateral support motor regions |
|  |  | 683 | p<0.001 | 7.19 | [-54;-12;33] | left postcentral gyrus |
|  |  | 280 | p<0.001 | 6.74 | [60;-9;33] | right postcentral gyrus |
|  |  | 68 | p<0.001 | 6.13 | [-3;-36;60] | left postcentral gyrus |
|  |  | 32 | p<0.001 | 6.06 | [12;54;39] | right superior medial frontal cortex |
|  |  | 119 | p<0.001 | 5.92 | [3;57;30] | right superior medial frontal cortex |
|  |  | 46 | p<0.001 | 5.48 | [-45;-69;-9] | left inferior occipital cortex |

Table S7. Regions showing significant differences in LCOR between individuals with lifetime depression and matched HC (Strategy Ⅱ)

| N criteria met | Contrast | Cluster size | Cluster p-value | Peak T statistic | MNI coordinates | Anatomical region |
| --- | --- | --- | --- | --- | --- | --- |
| 2 | DP<HC | 79 | p<0.001 | 6.22 | [27;-48;-51] | right inferior cerebellum |
|  |  | 38 | p<0.001 | 5.77 | [15;-45;-51] | right Lobule IX of cerebellar hemisphere |
|  |  | 259 | p<0.001 | 5.65 | [33;-84;-12] | right inferior occipital |
|  |  | 31 | p<0.001 | 5.24 | [0;-93;12] | calcarine |
|  |  | 31 | p<0.001 | 5.16 | [-42;-75;-9] | left inferior occipital |
| 3 | DP<HC | 2734 | p<0.001 | 6.94 | [-3;-30;60] | bilateral medial and superior sensorimotor regions |
|  |  | 190 | p<0.001 | 6.61 | [-12;-72;-51] | left Lobule VIII of cerebellar hemisphere |
|  |  | 168 | p<0.001 | 6.12 | [15;-60;-51] | right Lobule VIII of cerebellar hemisphere |
|  |  | 45 | p<0.001 | 6.12 | [-42;-6;3] | left insula |
|  |  | 50 | p<0.001 | 5.90 | [-12;-57;-48] | left Lobule IX of cerebellar hemisphere |
|  |  | 27 | p<0.001 | 5.76 | [-42;-51;-21] | left fusiform |
|  |  | 99 | p<0.001 | 5.54 | [-15;-48;-3] | left lingual and fusiform, parahippocampus |
|  |  | 27 | p<0.001 | 5.35 | [-39;-75;-12] | left fusiform |
| 4 | DP<HC | 623 | p<0.001 | 6.77 | [-42;-24;63] | bilateral medial and lateral sensorimotor regions |
|  |  | 172 | p<0.001 | 6.09 | [63;-15;30] | postcentral gyrus |
|  |  | 66 | p<0.001 | 5.58 | [0;-6;48] | left middle cingulate cortex |
|  |  | 64 | p<0.001 | 5.57 | [-15;-45;75] | left precuneus |
|  |  | 55 | p<0.001 | 5.56 | [51;-24;60] | right precentral gyrus |
|  |  | 38 | p<0.001 | 5.53 | [63;-9;-27] | right anterior middle temporal gyrus |
|  |  | 70 | p<0.001 | 5.52 | [-51;-66;-6] | left inferior temporal cortex |
| 5 | DP<HC | 92 | p<0.001 | 5.44 | [-45;-21;63] | left postcentral gyrus |

Table S8. Regions showing significant differences in GCOR between individuals with lifetime depression and matched HC (Strategy Ⅱ)

| N criteria met | Contrast | Cluster size | Cluster p-value | Peak T statistic | MNI coordinates | Anatomical region |
| --- | --- | --- | --- | --- | --- | --- |
| 3 | DP<HC | 42 | p<0.001 | 5.43 | [-24;-42;-3] | left lingual cortex and parahippocampus |

Table S9. Regions showing significant differences in GMV between individuals with lifetime depression and matched HC (Strategy Ⅱ)

| N criteria met | Contrast | Cluster size | Cluster *p*-value | Peak T statistic | MNI coordinates | Anatomical region |
| --- | --- | --- | --- | --- | --- | --- |
| 2 | DP>HC | 56 | 0.002 | 5.14 | [14;-33;75] | Right postcentral gyrus |
| 3 | DP<HC | 152 | 0.004 | 5.64 | [3;4;3] | Subcallosal cortex |
| 4 | DP<HC | 218 | 0.003 | 5.06 | [-44;-15;44] | left precentral gyrus |
|  |  | 64 | 0.015 | 4.87 | [0;-17;50] | left precentral gyrus |
|  |  | 62 | 0.015 | 4.69 | [0;32;-17] | Medial frontal cortex |
| 5 | DP<HC | 129 | 0.094 | 5.52 | [41;-24;48] | right postcentral cortex |
